# Investigating the evolution of undergraduate medical students’ perception and performance in relation to an innovative curriculum-based research module: a convergent mixed methods study launching the 8A-Model

**DOI:** 10.1101/2021.03.24.21254225

**Authors:** Farah Otaki, Deena AlHashmi, Amar Hassan Khamis, Aida Joseph Azar

## Abstract

Embedding an experiential research curriculum into medical programs is still not widely adopted. This study investigated, using convergent mixed methods design, the journey of medical students in relation to a research module. The students’ perception of the experience was qualitatively explored using thematic analysis. The students’ performance data were quantitatively analyzed using multi-repeated ANOVA.

The exploration generated four themes: 1-Attend-Acquire, 2-Accumulate-Assimilate, 3-Apply-Appreciate, and 4-Articulate-Affect. Quantitatively, two distinct clusters of mean Grade Point Average were revealed (p<0.01). Joint display analysis enabled integrating the qualitative and quantitative findings, generating the 8A-Model. The students start appreciating the experience upon conducting their research.

## Introduction

Early research training, particularly as part of undergraduate medical education, has been proven to significantly affect physicians’ career development (1-4) and professional identity (5-7). Physicians are more likely to perform research as one of their primary professional activities if they are exposed to research experiences early-on in their educational trajectory (8-10). For example, a study that investigated the impact of the Vanderbilt University School of Medicine Medical Scholars Program, showed that such programs prepare students for careers in academic medicine, and influences their career choices at an early juncture in their training (11). Such experiences are associated with improved academic performance, and increased research interest and productivity (2, 12).

The provision of quality medical care and the development of strong research skills, among healthcare practitioners, are inextricably linked (6). It is believed that research makes medical students better future clinicians and that it constitutes the core of the practice of medicine (1, 7). Research empowers the students to practice evidence-based medicine, enabling them to generate the necessary evidence to reinforce decisions during residency and in their future practice (7). In a study aimed at exploring the students’ perception of a unique student-driven Undergraduate Research Committee at Alfaisal University, Riyadh, Saudi Arabia, showed that such committees provide the future generation of physicians with diverse training opportunities to pursue research careers (3).

Embedding research curricula into undergraduate medical programs is still not widely adopted (2), although it is globally recognized to be an integral component of physician training (3, 13, 14). This is especially true if the learning opportunity is experiential in nature (15-18), and based on a holistic theory of education, which emphasizes learning as participation in the social world (19). To the best of the authors knowledge, there seems to be a gap in the literature regarding the trajectory through which undergraduate medical students go through to learn applied research concepts. Accordingly, this study aimed at investigating the journey of undergraduate medical students in relation to a compulsory curriculum-based research module, which has a prominent experiential component. The research questions of this study are as follows:

1. How do the MBBS students perceive the research module courses at different stages in their learning trajectory?
2. How is the performance of the students evolving as they progress in the module, and how does this trend relate to that of the students’ performance across all courses in the respective semesters?
3. What meta-inferences can be derived from integrating the qualitative data analysis (i.e., perception) with that of the quantitative one (i.e., performance)?

## Materials and methods

### Context of the study

This study was conducted at the Mohammed Bin Rashid University of Medicine and Health Sciences (MBRU) in Dubai, United Arab Emirates. The College of Medicine (CoM), at MBRU, offers an undergraduate Bachelor of Medicine and Bachelor of Surgery (MBBS) program (20).

This MBBS program consists of a six-year curriculum built on a competency-based learning model. The learning process is spiral, with integrated courses through-out the six-years, totaling 12 semesters (ie 2 semesters per academic year) (21). The CoM received its first MBBS batch of students in August 2016 (ie Class of 2022).

### Description of the research module under investigation

The research module is an integral part of the MBBS program at MBRU and is compulsory for all enrolled undergraduate medical students (22). Those students transition from secondary school directly into the program without academic induction. This module consists of a series of five interrelated courses in epidemiology, biostatistics, and research methodology delivered over the first five consecutive semesters of the MBBS program, Table 1. These 5-integrated courses are in complete alignment with the MBBS Program Learning Outcomes at MBRU. Where, upon completing those 5-integrated courses, the student would have developed the competences needed for the third Program Learning Outcome:

“*practicing evidence-based medicine, and engaging in scholarship and generation of new knowledge” (20)*.

**Table 1.**
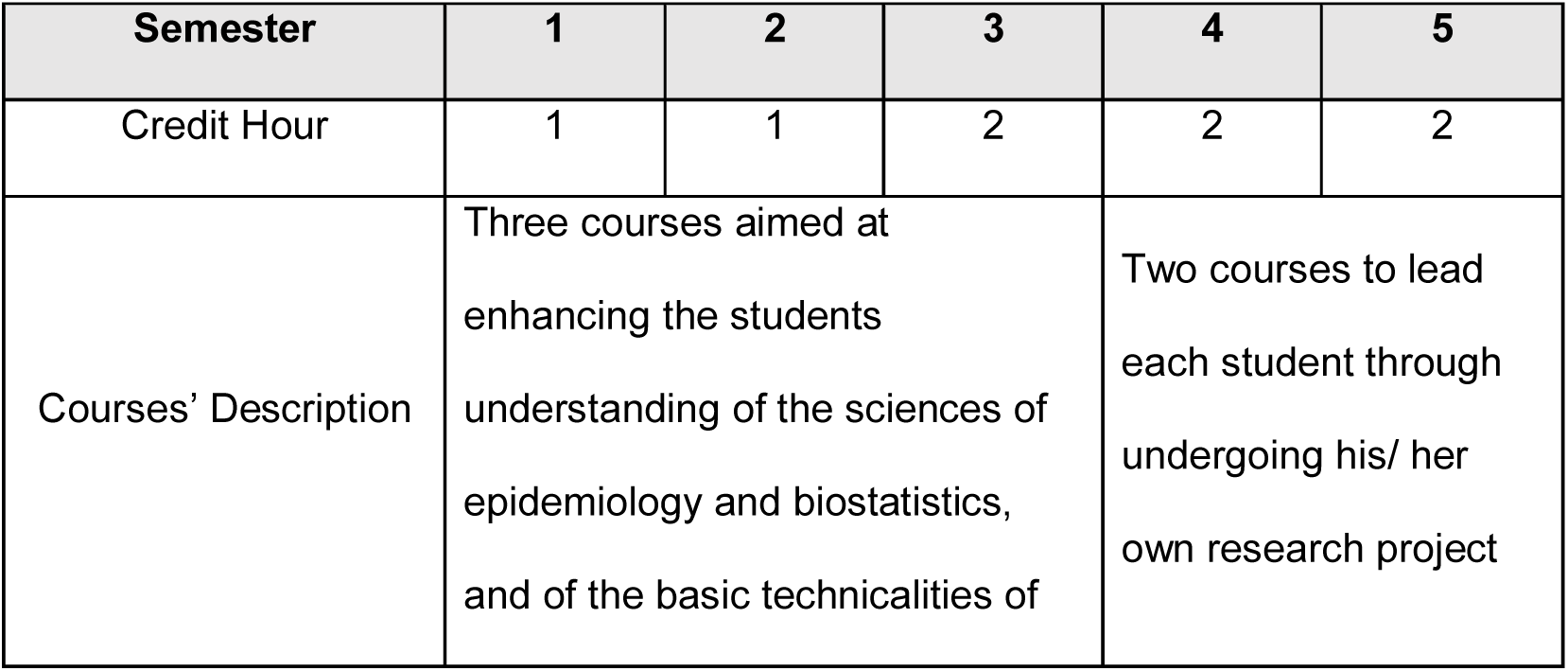
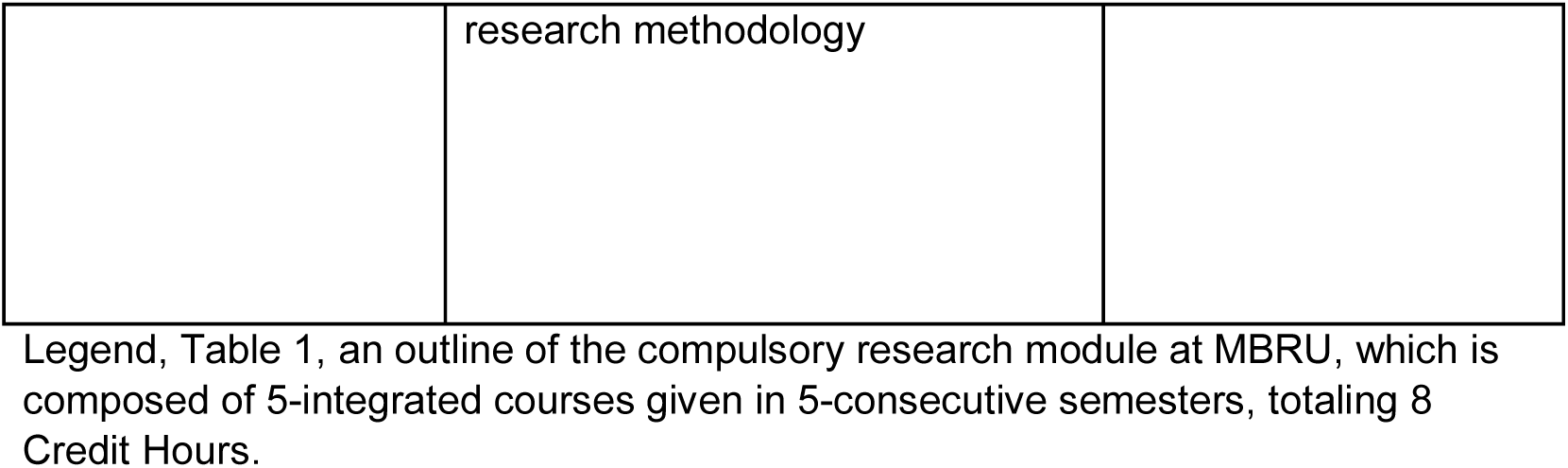
Illustrating the outline of the research module under investigation.

| Semester | 1 | 2 | 3 | 4 | 5 |
| --- | --- | --- | --- | --- | --- |
| Credit Hour | 1 | 1 | 2 | 2 | 2 |
| Courses' Description | Three courses aimed at enhancing the students understanding of the sciences of epidemiology and biostatistics, and of the basic technicalities of |  |  | Two courses to lead each student through undergoing his/ her own research project |  |

|  |  |
| --- | --- |
|  | research methodology |

Legend, Table 1, an outline of the compulsory research module at MBRU, which is composed of 5-integrated courses given in 5-consecutive semesters, totaling 8 Credit Hours.

Each course builds upon the knowledge and skills obtained in the preceding course. The mode of delivery is sequential that reinforces the acquired knowledge and skills, among the students, as they progress in the research module. Eight Credit Hours (CH) are awarded for completing this module: the two consecutive courses offered in semesters 1 and 2 are each awarded 1 CH. As for the courses offered consecutively in semesters 3, 4, and 5, each are awarded 2 CH, Table1.

The first three courses, delivered in Semesters 1 through 3 (totaling 4 CH), provide the students with a comprehensive background and understanding of the sciences of epidemiology and biostatistics, and of the basic technicalities of research methodology. The last two courses, delivered in Semesters 4 and 5 (totaling 4 CH), are more practical in nature, and are designed to further reinforce the students’ understanding of the principles of research design and methodology, where they apply what they learn through undergoing an actual research study, Table 1. This component of the learning experience is based on Situated Learning Theory, which is one of the holistic theories of education and emphasizes participation in the social world (19).

Each student performs their own research project, with an assigned supervisor, either basic or clinical sciences faculty member (depending on the scope of the study). The students are given an extensive list of health-related research opportunities of which they are given the autonomy to choose from. These projects are biomedical (clinical or lab-based) or socio-behavioral in nature; the projects in the latter track are related to health systems or medical education. The supervisors’ main responsibility is to co-create with their assigned supervisees, while mentoring them in relation to the subject matter throughout the scientific research process. As such, the individual students go through a process of adaption as they progress in the learning experience, which takes place in the authentic context, among the respective community of practice; all of which depends on what the individual student’s research project is.

The course learning objectives guide the instructional learning strategy of each course. The first three courses are delivered using both lecture-based teaching and case-based learning. The summative assessment contributes to the Grade Point Average (GPA) which is out of 4.0 and constitutes a direct measure of the individual students’ performance. The attainment of the learning objectives of these courses is measured by a combination of summative and formative assessments: Assignments (formative), Open-book Assessment (summative), and End-of-term Examination (summative). The course instructors are responsible for grading all the assessments.

As for the last two courses, they are delivered using both case-based teaching and project-based learning. The attainment of the learning objectives in semester 4 is measured by a combination of summative and formative assessments: Research Ethics Module (formative), Student Research Project Form (formative), Research Proposal (summative), In-course Assessment (summative), Student Progress (summative), and Oral Presentation (summative). As for semester 5, students are assessed on the following parameters: Conference Poster Presentation (summative), Research Project Dissertation (summative), Student Progress (summative), and Digital Abstract (summative). It is worth noting that the same group of instructors (three faculty members) are involved in delivering the 5-integrated courses and in grading all entailed assessments (using pre-defined rubric). The assessment plan, question type, and grading method used are congruent across all cohorts. Accordingly, consistency of assessment and corresponding grading scheme is ensured.

### Research design

A convergent mixed methods study design was adapted to develop a systemic understanding of the experience of the undergraduate medical students throughout the respective research module. For that matter, qualitative and quantitative datasets were concurrently collected and in turn systematically integrated. The triangulation of data, as such, enables investigating the same phenomenon from differing perspectives, which in turn raises the validity of the generated findings.

The qualitative segment of this study aimed at exploring the development of the perception of students in relation to the module, and to its immediate output, and foreseen outcome and long-term impact. It relied on a phenomenological research methodology (23, 24) based on the deployment of the Braun and Clarke (2006) six-step model of conducting thematic analysis (25). This framework, which is based on a constructivist epistemology, is commonly used in the realm of social and behavioral research (26, 27).

In parallel, the performance of the students as they progress through each course of the research module was quantitatively analyzed. This quantitative segment relied on a cross-sectional time-series design (28), using the GPA of the students of all three classes (2022, 2023, and 2024) on an aggregate level. The performance of the students in each of the courses of the research module was measured. The generated trend across those courses was compared to that of the students’ performance of all courses (of the MBBS program) in the respective semesters.

### Ethical Approval

Ethical approval for the study was granted by the MBRU, Institutional Review Board (Reference # MBRU-IRB-2020-015).

### Participants

The study was conducted in Fall 2019-2020 and involved three cohorts of MBBS students: Class of 2022 who had completed the entire research module, all 5 integrated courses (8 CH), Class of 2023 who had completed the first four courses (6 CH) out of 5 courses, and Class of 2024 who had completed the first two courses (2 CH) out of 5 courses, Table 2.

**Table 2.**
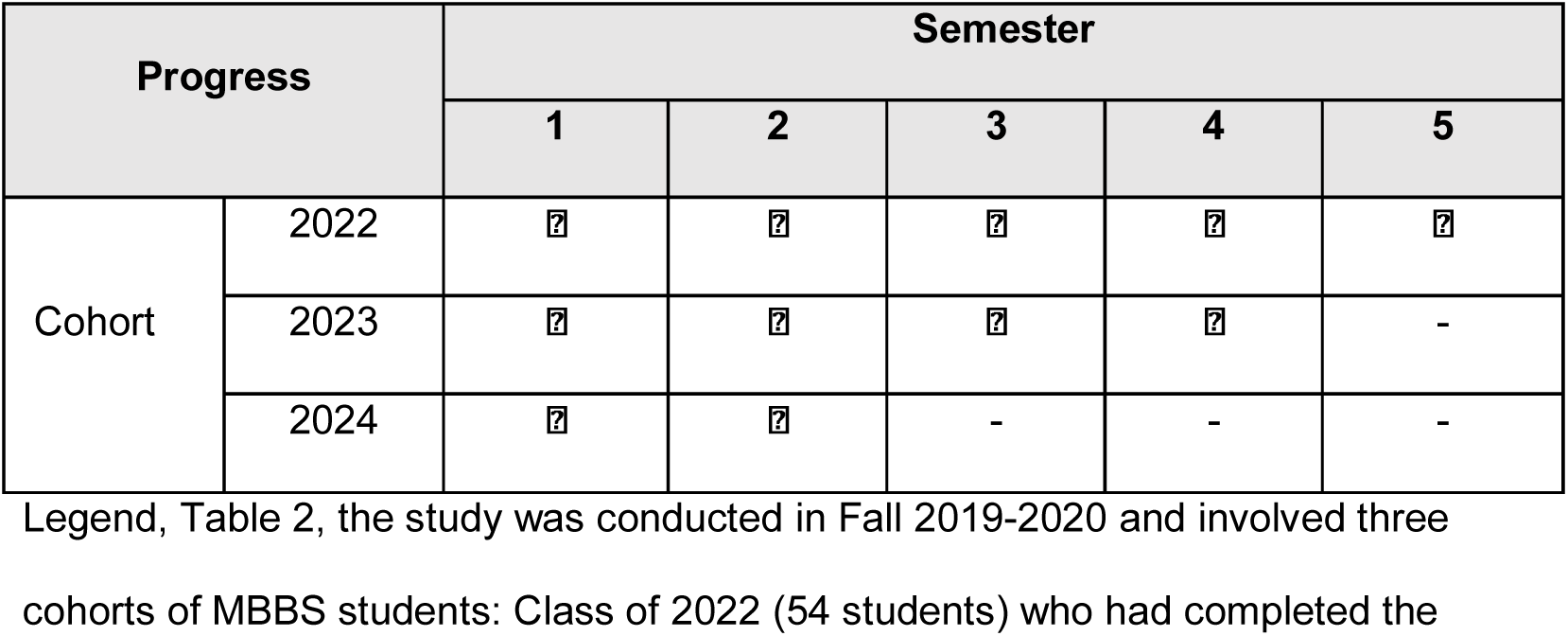
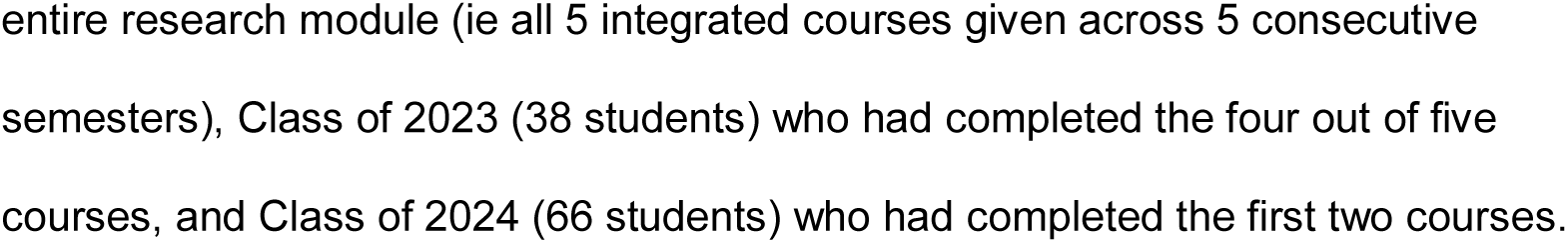
Illustrating the respective cohorts progress in the research module under investigation.

## Data Collection

### Qualitative data

The qualitative segment of this study involved exploring the evolution of the perception of the students regarding their experiences as part of the research module from joining the MBBS program. For that matter, three focus group sessions were conducted. Since the ideal size of a focus group is between five to eight participants (29, 30), seven randomly selected students, from each of the three classes (2022, 2023, and 2024), were invited to the focus group sessions (one corresponding to each class). The randomization was done using the ‘Select Cases’ function of SPSS statistical package (Windows version 25.0, IBM Corp, NY, USA).

A focus group protocol was tailor-made to guide the data collection initiative across all three sessions. Forty-five minutes were allocated for each group session, which was divided into four segments. The first 15 minutes inquired about the knowledge, skills, and competencies acquired through the module (including but not limited to: basic concepts and research process). The following 10 minutes were focused on the research learning experience, across the module, and the output, outcome, and impact of the module. The next 10 minutes included discussions around the effectiveness of the module, the students’ level of appreciation, and the value of research. The last 10 minutes constituted the wrap-up where the strengths and opportunities for improvement of the module were pinpointed. These focus group sessions were facilitated by a researcher experienced in designing and undergoing qualitative studies (FO).

The data collection tool, which was designed especially for this study, underwent two validation phases. Firstly, five faculty members at CoM were contacted for the content validity. Secondly, the questions of the tool were discussed with 10 randomly selected students to assess the readability and comprehensibility of the questions, and the sequence by which the questions were presented (ie face validity).

Students were informed that participation in the focus group sessions was completely voluntary and were given the option to withdraw from participation any time before or during the focus group session. In addition, the students were requested to provide written consent as a prerequisite to participation. Participating students were assured regarding the anonymity and confidentiality of the data generated from the respective sessions.

Each participant was given a unique identification number (ie participants were numbered 1 through 15). These unique identification numbers were complimented with ‘1’ for Class of 2024, ‘2’ for Class of 2023, and ‘3’ for Class of 2022 (ie participants 1 through 6 were followed by ‘1’, 7 through 11 by ‘2’, and 12 through 15 by ‘3’).

### Quantitative data

Assessment data, including but not limited to the GPA, is routinely gathered by the Student Services and Registration department at MBRU (SSR). Data on the students’ performance (GPA), per semester, across all courses offered in the respective semester, and in relation to each of the sequential courses of the research module were requested by the researchers from the SSR. The obtained GPA values corresponded to Semesters 1 through 5 (Class of 2022), Semesters 1 through 3 (Class of 2023), and Semester 1 (Class of 2024). This depended on where the respective students were in their learning trajectory. All the data related to the performance of the students was deidentified by a member of the SSR department prior handing it over to the research team.

## Data Analysis

### Qualitative data

The qualitative data was thematically analyzed by three researchers (AJA, DA, & AH), following the six steps of the abovementioned framework (25). The data collected from each of the three cohorts was handled separately; the researchers were not informed which cohort each of the datasets corresponded to. The researchers started with familiarizing themselves with the data; patterns across the datasets were systematically identified and reflected upon. The second step included segmenting the data into meaningful statements and generating initial codes. The third step included searching for themes. This led to the generation of a set of themes that refer to differing stages of the students’ learning journey. The themes were then reviewed as the fourth step, where they underwent several rounds of reflections.

The three researchers, as part of the fifth step, agreed on the optimal way to sequence those themes based on their collective understanding of the encapsulating context, delivered curriculum, and receiving students. They also coded the themes. The researchers factored into the analysis their interpretation of differing knowledge transfer, exchange, and valorization theories (eg Kirkpatrick model, Learning-Transfer Evaluation, system thinking, knowledge management, and processual analyses) (31-36).

NVivo software version 12 plus (QSR International Pty Ltd, Vic, Australia) was used to code the data, as per the agreed upon conceptual framework, and in turn facilitate the categorization of the relevant text fragments. The transcripts were examined, line-by-line, while coding the text fragments that relate to the research questions until no new information was observed in the data, and hence data saturation was attained. All this paved the way for the last step of the adapted framework which constituted the basis of reporting upon the results and was done in alignment with recently published recommendations for reporting qualitative research (37, 38).

### Quantitative data

The extracted quantitative data was analyzed using the SPSS statistical package (Windows version 25.0, IBM Corp, NY, USA). Continuous data was described by measures of tendency and dispersion. Analysis of variance was used to compare the mean GPA in the different semesters. A p-value of less than 0.05 was considered statistically significant (39).

### Mixed Methods Integration

The findings from both types of analyses (qualitative and quantitative) were mapped onto each other and carefully reflected upon. This mixed methods integration took the form of an iterative process, namely: the joint display analysis (40). This ultimately led to meta inferences (41). The researchers explored how the output of the analyses relate to one another to synthesize a meaningful narrative.

## Results

### Qualitative

Out of twenty-one randomly selected students who were invited to the sessions, 15 (71%) agreed to participate: 4 (Class of 2022), 5 (Class of 2023), and 6 (Class of 2024).

The generated data included reflections of the research learning trajectory, from the perception of the students. As illustrated in the conceptual framework generated as part of this study (Fig 1), the thematic analysis depicts four sequential steps, namely: 1-Attend-Acquire, 2-Accumulate-Assimilate, 3-Apply-Appreciate, and 4-Articulate-Affect, that the students go through as part of the research module to effectively integrate the scientific research method, Table 3.

**Table 3.**
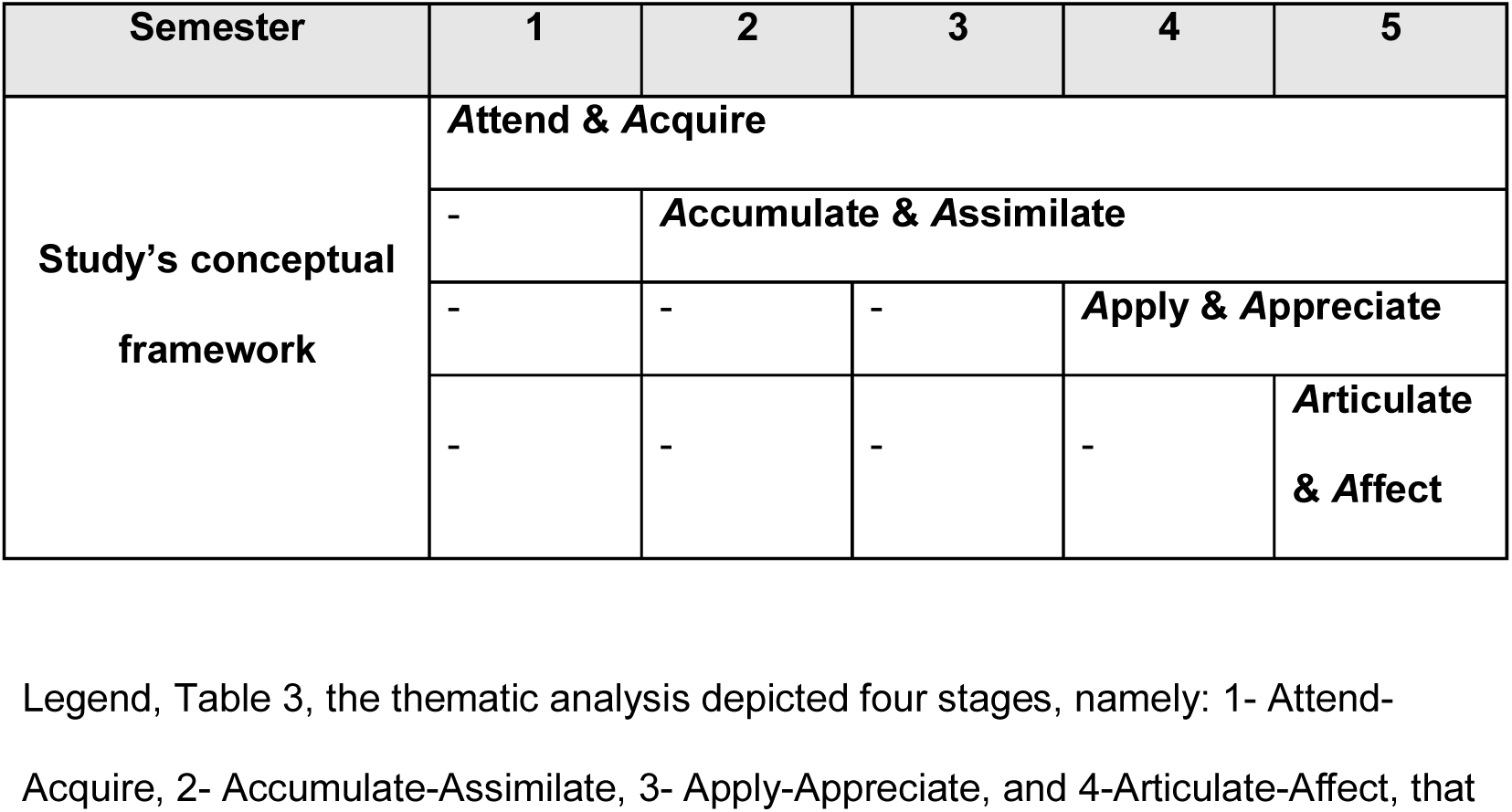
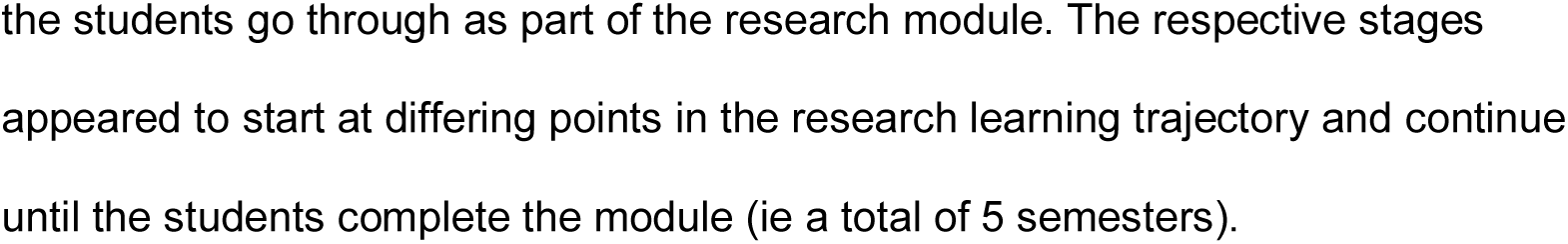
Mapping the study’s conceptual framework (output of analysis of the qualitative component of this study) onto the courses of the research module under investigation.

**Fig 1.**
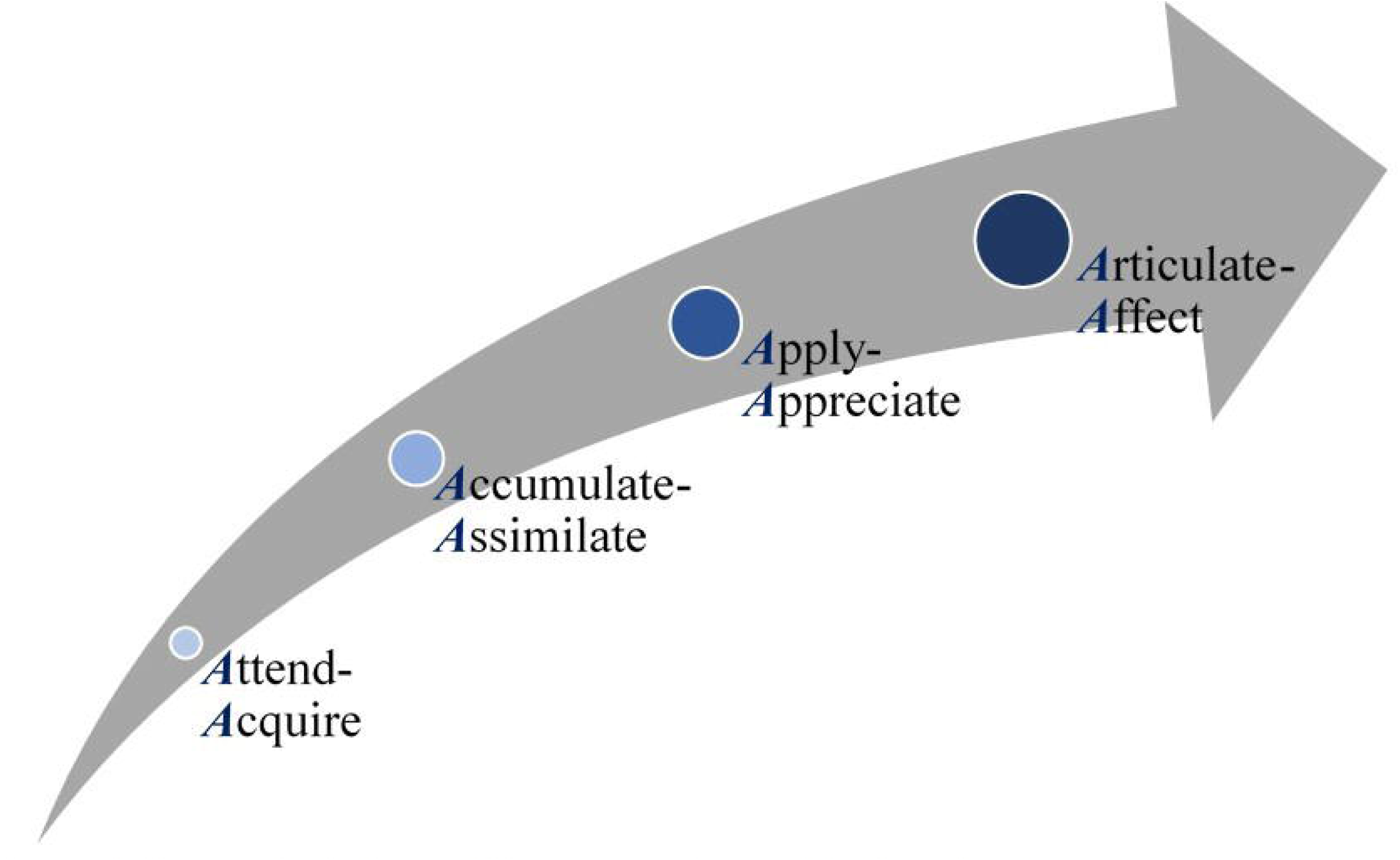
Sequential steps of the research module learning journey.

Fig 1 legend, the qualitative component of this mixed methods research study generated this framework, depicting the different phases that students go through as part of the curriculum-based research module (represented as the Grey arrow). Each phase, representing a theme of the inductive analysis, appeared to be characterized by an interplay of two verbs. Hence, the code mindfully ascribed to each theme brings together two verbs, each starting with the letter ‘A’. The increasing size of circles represent the students’ accumulation of expertise (ie integrated knowledge and skills), and the color development (ie from light to dark) represents the gradual evolution that the students go through, over time.

### Attend-Acquire

The first theme encapsulated the text fragments that refer to the attendance of the students, which highlighted the students’ enrolment, their in-class experience and contributions, and their interactions with the instructors. The content of the first two courses was described as novel, and in some cases, confusing, to those who had just started their journey.

> 1.1. “…before we start the lecture, I go over the learning objectives, and as the instructor goes through the presentation explaining its content, I would be ticking off the learning objectives to confirm that we covered them all…”
>
> 6.1. “…while the instructors are delivering the biostatistics content, we continuously shift between the theory and the practice using SPSS. It is difficult for us to see the link. We do not see how the theory translates into practice. We are told there are links, but we do not see them. This confuses us…”

This theme also included text fragments that related to the knowledge and skills that the students acquired from their experience.

> 2.1. “…we learned about the different types of studies. For example: experimental and non-experimental studies. We will be using this knowledge to design our own research studies at a later stage…for biostatistics, it is all based on formulas. We need to practice it step-by-step to be able to understand and implement it in our research studies…”
>
> 5.1. “…we learned about public health… it is the art and science revolving around the health of the population…it shows you how it is all interlinked…the health and wellbeing of one patient is related to the status of the community that s/he belongs to…”
>
> 7.2. “… we learned mostly about research. We started off the module learning about epidemiology… In parallel, they were teaching us biostatistics. We did not know back then the relevance and importance of biostatistics. Its true importance became apparent to us in Semester 4…”
>
> 9.2. “…I am now better at writing professional emails, because there are so many emails that I had to send to my supervisors; there were a lot of correspondences, back and forth, between us…”

### Accumulate-Assimilate

The second theme is related to how the knowledge and skills are accumulating, and in turn shaping the students’ attitudes and habits (eg critical appraisal and retrieving evidence). At this stage in the learning curve, the students start realizing the importance of the disciplines of biostatistics, epidemiology, and research, and how they are interlinked:

> 3.1. “…this course enabled us to understand articles. We critically appraise articles. We gained the habit of screening data, information, and knowledge. We evaluate the quality of the evidence prior taking it into account…”
>
> 7.2. “… we now know how to properly read a peer-reviewed article. We do not just read the findings from the perception of the investigators. We investigate the numbers, the calculations, and the statistical techniques. We check the reliability of the study and the validity of the results. We critically-appraise its core, along with going through and reflecting upon the findings of the authors…”
>
> 8.2. “… in the beginning, we were given epidemiology and biostatistics. Back then, I did not understand neither their importance, nor their relevance to each other…I started seeing the link between them in the fourth semester…”
>
> 14.3. “… it is important to know how to effectively interpret data if one wants to implement evidence-based medicine…”
>
> 15.3. “…I started to use what I acquired from this module in other courses…the five courses turned out to be interlinked. …”

Then comes the stage, where the students seem to play an active role in assimilating, and in turn integrating the acquired skills and knowledge. They start building expertise and resilience:

> 1.1. “…knowledge and skills of research are very important, and go hand-in-hand with practicing medicine… as clinicians, we need to continue on reading articles, keeping an eye on new studies so we can stay up-to-date…”
>
> 11.2. “… my supervisor effectively mentored me to integrate all that I had been acquiring to ask the right questions. It was not an easy journey for me, but I made it through. It has been so enriching…”

### Apply-Appreciate

Following that, the students start applying what they have been acquiring, accumulating, and assembling. They feel empowered and a sense of ownership while undergoing their own research study:

> 7.2. “…for you to know something very well, you would have to do it…”
>
> 9.2. “…I used to always hear about research and physicians doing research, and that would get me really excited, but I did not know what research is all about…Now, I am informed, and feel empowered to attain my aspirations…”

After the students start putting into practice what they have been learning, they start appreciating the journey and all that it entails. The students have expressed gratitude, and in some cases-excitement:

> 9.2. “…we are grateful that we are given this opportunity at an early stage in our educational journey…”
>
> 12.3. “…my research topic was a literature review. I am now proficient in running systematic literature reviews. This is helping me during my internships, and as I am studying for other courses…”
>
> 14.3. “… this module made me appreciate research and how research is done. I really enjoy the scientific research process. I felt a strong sense of accomplishment upon completing and submitting my research study…I am now involving myself in other research opportunities…”

### Articulate-Affect

The last theme included reflections in regard to how the students are articulating their findings and contributing to the theory (and practice) of the subject matter (be it through presenting and/ or publishing), which they are convinced would enrich their professional profiles.

> 9.2. “…we get a lot of opportunities to share, present, and collectively reflect upon our work…it is good to indicate this experience on our professional profiles…”
>
> 13.3. “…I learned what it takes to publish an article, and that, as they say: ‘the whole is more than the sum of its parts’…”
>
> 14.3. “…I learned how to design…I designed a research study and wrapped-up the experience with designing a poster presentation and a conference digital abstract video. You need to learn how to ask the right questions, and to be equipped with the knowledge and skills to answer them effectively…”

This theme also related to how the students perceive themselves to be affecting and altering the field by practicing evidence-based medicine, improving performance (clinical or otherwise), and developing communities. They also refer to how they are leveraging the expertise that they acquired in this module to coach others:

> 7.2. “…this is particularly relevant to our region. By conducting research and generating knowledge, we will be contributing to the development of our countries…”
>
> 13.3. “… this module enabled me to understand the importance of research and practicing evidence-based medicine…”
>
> 14.3. “…I learned a lot from the entire research experience…I am using what I gained from these courses in my internships…”

## Quantitative

### Sample description

The total number of students in all 3 classes (2022, 2023, and 2024), whose GPA values were factored into the analyses, was 158. Yet, the number of students per semester differed as this was dependent, by design of the study, on where the respective students were in their learning trajectory: Class of 2022 (54 students) who had completed the entire research module; Class of 2023 (38 students) who completed the first 4 Semesters; and Class of 2024 (66 students) who completed the first two semesters. In other words, 158 students completed Semesters 1 and 2, 92 completed Semesters 3 and 4, and 54 completed Semester 5.

### Research module

The quantitative analysis resulted in two distinct clusters of mean GPA values across the 5-integrated courses’ module given in semesters 1 through 5, as shown in Fig 2. The first cluster was for courses delivered in semesters 1, 2, and 3 where the mean GPA values were found to be homogenous (p>0.05). The second cluster, which was also established to be homogeneous (p>0.05), was for the mean GPA values of students across the courses offered in semesters 4 and 5. Moreover, the mean GPA values for Semesters 1, 2, and 3, as compared to Semester 4 and 5, was significantly lower in the former cluster (p<0.01). The same trend, with a similar significance, was observed when the data of Semester 1 to 5 of Class of 2022 only was analyzed. It became evident that two significantly different clusters were observed cluster 1 (Semesters 1, 2, and 3) and cluster 2 (Semesters 4 and 5) (P<0.01).

**Fig 2.**
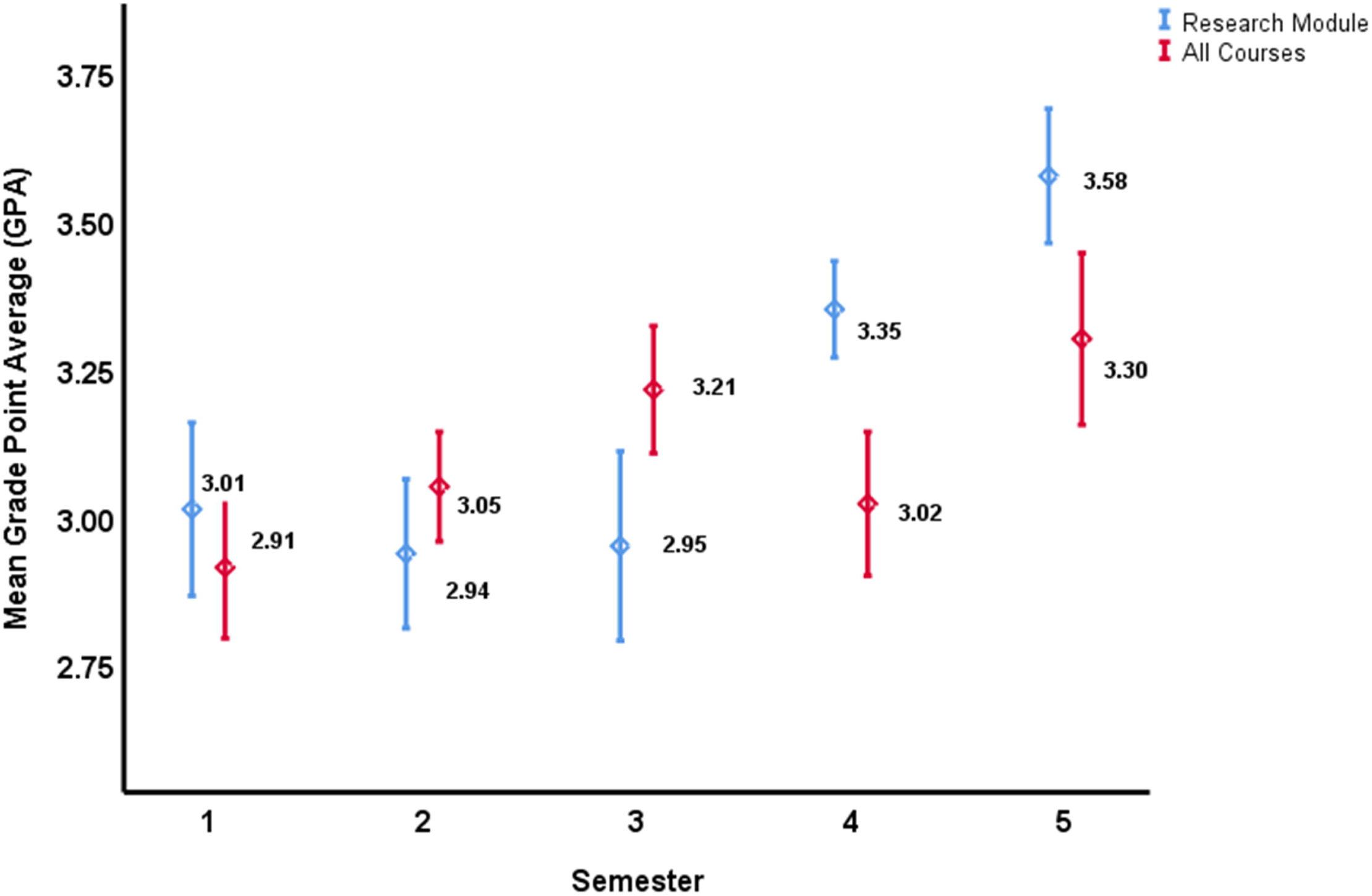
Mean Grade Point Average (GPA), with the corresponding 95% Confidence Interval (CI), for the research module courses (Blue), and all the MBBS program courses together (Red), offered in the respective *semester*.

Fig 2 legend, Blue lines-There are two distinct clusters of mean GPA values across the 5-integrated course modules: first cluster was for courses of semesters 1, 2, and 3, and the second cluster was for courses of semesters 4 and 5. The 95% CI for Semester 1, 2, and 3 overlap indicating no statistical difference between the three research courses given in the first 3 semesters. Similarly, the 95% CI for the research courses given in Semester 4 and 5 overlap indicating no statistical difference (p>0.05). However, post-hoc ANOVA analysis revealed two distinct clusters of mean GPA, the first cluster (Semesters 1, 2, and 3) and the second cluster (Semesters 4 and 5) (p<0.01).

### All program courses

As evident in Fig 2, the mean GPA values of the students, across all courses offered as part of the MBBS program (including the courses of the research module), was heterogeneous. The results showed that the mean GPA in semester 1 was significantly different than that of semesters 2, 3, and 5 (p<0.01); semester 2 was significantly different than that of semesters 1 and 5 (p<0.01); semester 3 was significantly different than that of semesters 1 and 4 (p<0.01); semester 4 was significantly different than that of semesters 1 and 2 (p<0.01); and semester 5 was significantly different than that of semester 3 (p<0.01). Indicating the absence of any clustering.

## Mixed methods integration

Merging the output of the thematic analysis into that of the quantitative analysis unearthed a holistic perspective of the situation, illustrated in the study’s joint display (Fig 3). The convergence of findings enabled the development of a thorough understanding of how the students’ perception and performance evolve as they progress through the module. On its own, the narrative analysis showed the 4 steps that the students go through as part of the journey, with particular emphasis on the value of the step where the students start putting into practice what they have been learning. As for the quantitative analysis (on its own), it revealed that the students’ performance can be observed as two distinct clusters. These findings confirmed the results of the qualitative segment of this study, illustrating that the turning point in the learning trajectory of the students is at the “Apply” step of the introduced conceptual framework.

**Fig 3.**
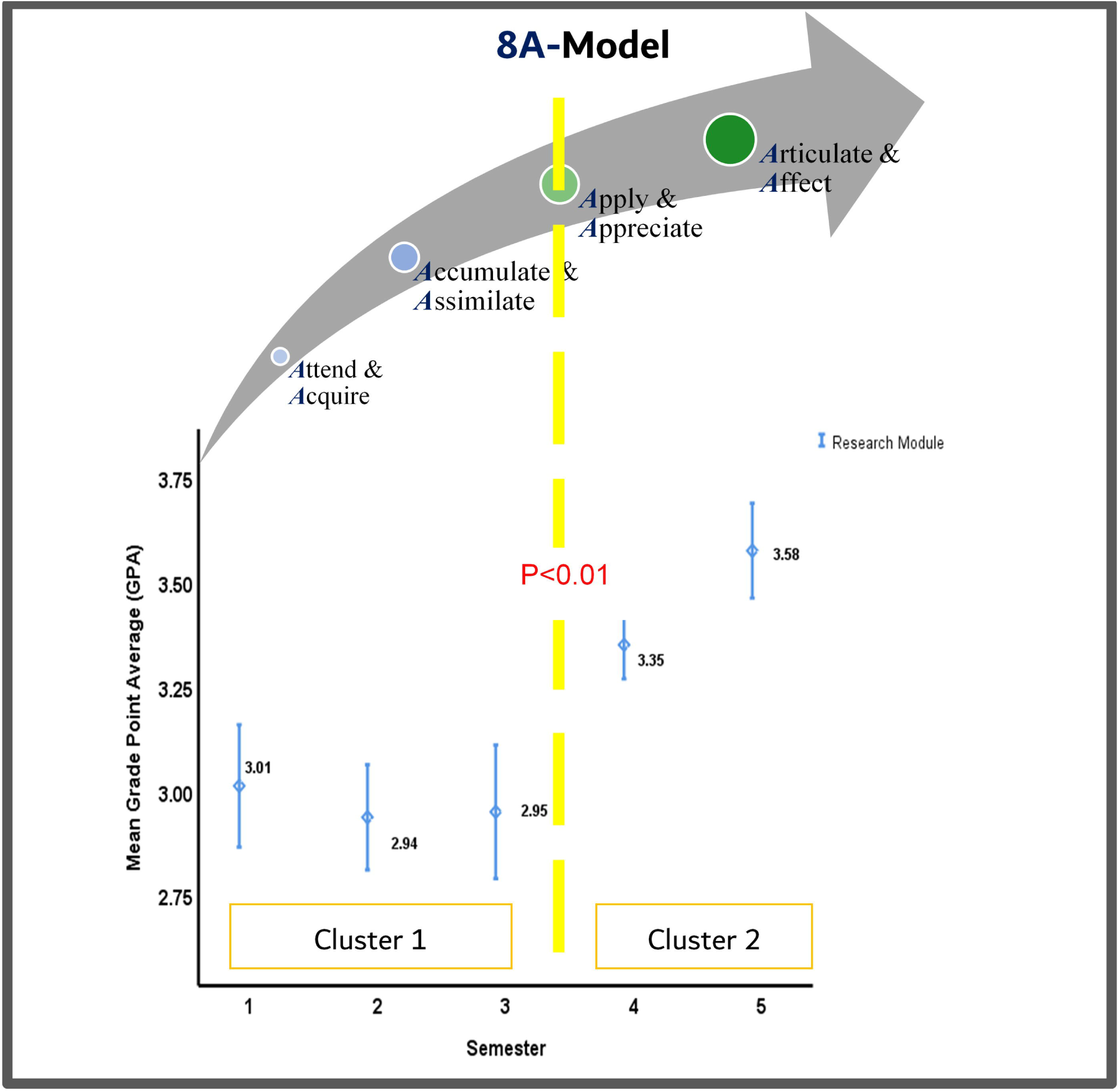
A joint display, mapping the qualitative findings (the study’s conceptual framework) with the quantitative analysis (dispersion of cluster-level performance measure) in the 5-integrated research courses across 5 consecutive semesters (semesters 1, 2, and 3 where students were ‘taught about research’ and semesters 4 and 5 where students were ‘enabled to conduct research’).

Fig 3 legend, mapping the study’s conceptual framework onto the student’s performance revealed a holistic perspective of the situation. The convergence of findings, as such, confirmed the existence of a turning point (highlighted with a Yellow dotted line). The students started expressing appreciation of the value of the learning experience upon putting into practice the knowledge and skills that they were acquiring, accumulating, and assimilating. The color transformation [ie from a primary color (ie Blue) to a secondary color (ie Green: Blue + Yellow)] further emphasizes this turning point, and how the increased appreciation happened concurrently with their increased engagement and enhanced performance.

## Discussion

This study introduced the evidence-driven 8A-Model. This framework, in alignment with the Kirkpatrick’s four level model (42, 43) and other theories around knowledge acquisition (44-46), suggests that the undergraduate medical students enrolled in an integrated research module go through specific steps in order to effectively integrate the scientific research method. These steps start with students attending the courses, followed by acquiring and in turn accumulating the knowledge and skills. Next the students assimilate what they are acquiring, which in turn, enables them to apply what they have been learning all along. This empowers the students, solicits their appreciation, and encourages them to articulate their findings, which in turn affect the fields of medicine and/ or public health, instilling constructive change and improvements.

In parallel, the quantitative analysis revealed that the learning curve integral to the trajectory of the students, as part of the research module, can be observed as two segments. From the first through the third course, the three averages of performance of the students were similar. The averages of performance significantly increased in the fourth and fifth courses. Congruent with the results of the qualitative segment of this study, the turning point is at the “Apply” step of the introduced framework. As suggested, in previous studies, it is not enough to teach students about research (47). Students need to be provided with the opportunities to conduct research, and to be counselled on the attitudes needed for them to thrive in research environments (47, 48). This is especially true when, similar to the case of the research module under investigation in this study, the experiential learning component is based on the Situated Learning Theory (19, 49), and when the educator considers individuals, and their experiences and environments (50, 51). As such, the experiential education was maximized through the individual students’ embeddedness in the authentic context, among a community-of-practice. Their active adaption is facilitated by experts in the subject matter.

Along these lines, this study showed that the students start to truly appreciate the value of the learning experience upon using the knowledge and skills that they are acquiring, accumulating, and assimilating. This appreciation is reflected in their increased engagement and enhanced performance. It is worth noting that this observation is quite different than the suggestions of other traditional knowledge transfer and integration models (eg Kirkpatrick model and the Learning-Transfer Evaluation) which highlight appreciation (or the lack of it) as an instant result of participating in any one learning experience (32, 43). When it comes to effectively integrating the scientific research method, there is a prominent learning curve with a gradual evolution, and the content acquired is usually novel. So, it takes time to properly digest the acquired content, and start putting it into practice.

Moreover, the results show that the performance of the students throughout the research module is more consistent, with less fluctuations, than their performance across all courses in the first five semesters of the MBBS program.

This could be indicative of the level of integration within each course of the research module, and across all five courses of the respective module. It is worth highlighting that although the 8A-Model is diagrammatically depicted as a linear process. It is more likely to be an iterative one given the required spiral mode of delivery which includes purposeful repetition of specific content to ensure effective integration within each course and across all five courses.

The convergent mixed methods study design enabled the development of thorough insights into this innovative research module, and its application as part of an undergraduate medical program. Yet, the generalizability of the results is limited to contexts that are like MBRU. Hence, it is worthwhile for future studies to investigate the application of such modules, and the validity and reliability of the generated 8A-Model across multiple medical programs. Moreover, by virtue of the selected study design no causality can be established. It would be worthwhile for future studies to investigate the same variables (ie perception and performance) longitudinally (preferably through an experimental design) to better understand how the respective variables relate to one another.

## Conclusion

The evidence-driven 8A-Model, generated by this study, highlights that students’ understanding of the true value of research seems to increase as they progress in the module. They begin expressing appreciation of the significance of the experience when they start implementing what they are learning (acquiring, accumulating, and assimilating) as part of their own research studies. It is recommended for such a research module, with a firm experiential component, to be integral to undergraduate medical programs.

## Data Availability

All data referred to in the manuscript can be obtained from the corresponding author upon request.

## Acknowledgement

The corresponding author would like to extend gratitude to Dr. Tom Loney for his valuable contribution to delivering components of the research module under investigation as part of this study.

## Supporting information

### Glossary terms and abbreviations

8A-Model: The framework generated by this study to suggest that students go through four phases of an iterative process: *A*ttend-*A*cquire, *A*ccumulate-*A*ssimilate, *A*pply-*A*ppreciate, and *A*rticulate-*A*ffect, with an evident turning point around “Apply”.

CoM: College of Medicine
CH: Credit Hours
GPA: Grade Point Average
IRB: Institutional Review Board
ANOVA: Analysis of Variance
MBBS: Bachelor of Medicine, Bachelor of Surgery, or in Latin:Medicinae Baccalaureus, Baccalaureus Chirurgiae
MBRU: Mohammed Bin Rashid University of Medicine and Health Sciences
SD: Standard Deviation
SPSS: Statistical Package for the Social Sciences
UAE: United Arab Emirates

## Ethics approval and consent

Prior to the commencement of the study, written informed consent was obtained from all participants. Ethical approval was obtained from Ethical approval for the study was granted by the MBRU, Institutional Review Board (Reference # MBRU-IRB-2020-015).

## Notes on contributors

FO-collected and analyzed the data, designed the 8A-Model with inputs from all authors, composed the final version of the manuscript; DAH-helped in analyzing the data and curation, and assisted in revising the manuscript; AHK-contributed to delivering components of the research module, and analyzed the data and assisted in revising the manuscript; AJA-developed and facilitated the delivery of the research module, maintained the macro perspective of the study, drafted the protocol, analyzed the data, composed, approved and submitted the final submission of the manuscript with inputs from all authors. All authors approved the final manuscript.

## Availability of Data

The dataset is not publicly available but can be shared by the corresponding author (AJA) upon request.

